# Factors associated with initiation of bone-health medication among older adults in primary care in Ireland

**DOI:** 10.1101/2020.10.05.20207233

**Authors:** Mary E. Walsh, Mari Nerdrum, Tom Fahey, Frank Moriarty

## Abstract

**Background:** Adults at high risk of fragility fracture should be offered pharmacological treatment when not contraindicated, however under-treatment is common.

**Objective:** This study aimed to investigate factors associated with bone-health medication initiation in older patients attending primary care.

**Design:** Retrospective cohort study.

**Setting:** 44 general practices in Ireland from 2011-2017.

**Subjects:** Adults aged ≥65 years who were naïve to bone-health medication for 12 months.

**Methods:** Overall fracture-risk (based on QFracture) and individual fracture-risk factors were described for patients initiated and not initiated onto medication and compared using generalised linear model regression with Poisson distribution.

**Results:** Of 36,799 patients (51 % female, mean age 75.4 (SD=8.4)) included, 8% (n=2,992) were observed to initiate on bone-health medication during the study. One fifth of all patients (n=8,193) had osteoporosis or had high fracture-risk but only 21% of them (n=1,687) initiated on medication. Female sex, older age, state-funded health cover and osteoporosis were associated with initiation. Independently of osteoporosis and co-variates, high 5-year QFracture risk for hip (IRR=1.33 (95% CI=1.17-1.50), p<0.01) and all fractures (IRR=1.30 (95% CI=1.17-1.44), p<0.01) were associated with medication initiation. Previous fracture, rheumatoid arthritis, and corticosteroid use were associated with initiation, while liver, kidney, cardiovascular disease, diabetes and oestrogen-only hormone replacement therapy showed an inverse association.

**Conclusions:** Bone-health medication initiation is targeted at patients at higher fracture-risk but much potential under-treatment remains, particularly in those >80 years and with co-morbidities. This may reflect clinical uncertainty in older multimorbid patients, and further research should explore decision-making in preventive bone medication prescribing.

**Key points:**

- During this study, 23% of women and 11% of men defined as high-risk of fracture were newly initiated on bone-health medication.
- Rates of potential under-treatment were highest in patients over 80 years old.
- Fracture history, corticosteroid use, and rheumatoid arthritis were independently associated with medication initiation.
- Patients with diabetes, and liver, kidney, or cardiovascular disease were less likely to be initiated on medication.
- Clinical guidelines should provide advice on risk-benefit decisions in osteoporosis treatment where co-morbidities are present.

## INTRODUCTION

Fragility fractures, usually caused by falls in the presence of osteoporosis, are a prominent cause of morbidity and mortality, affecting approximately 3% of older adults annually [1,2]. Hip fractures are particularly serious with a 20% one-year mortality rate [3].

International guidelines recommend that adults at high fracture-risk, including those with osteoporosis, previous fractures or taking long-term corticosteroids, be offered pharmacological treatment where no contraindication exists [4-6]. There is strong evidence that oral bisphosphonates reduce fracture incidence in men and women and they are the first line-therapeutic choice [4,7,8]. A common alternative treatment denosumab, which reduces fracture incidence in women and improves bone mass density (BMD) in men, involves six-monthly administration by sub-cutaneous injection [9-11].

It is well recorded that osteoporosis is under-diagnosed and under-treated. A nationally representative Irish study showed that 12% of women and 3% of men aged over 50 years had objective osteoporosis but only 28% of them were diagnosed [12]. Even in those diagnosed with osteoporosis, or identified as high fracture-risk, pharmacological treatment is not initiated in 23-72% of patients [9,13]. Reasons for non-initiation include GP and patient concerns about medication side effects (particularly gastrointestinal), perceived uncertainty about effectiveness, medicine administration restrictions and costs [9,13,14].

In primary care, it is recommended that fracture-risk be determined by risk prediction tools including FRAX and QFracture, which can be supplemented by BMD assessment [4,15,16]. Common factors included in these tools are older age, female sex, previous fracture, medications and conditions that affect bone quality and disorders that increase falls-risk [15,16]. Identification of factors associated with new initiation of bone-health medication in a large primary care population could reveal cases in which osteoporosis and high fracture-risk is potentially under-identified or under-treated in the Irish setting.

This study aims to investigate if older patients at higher fragility fracture risk had higher rates of new initiation of bone-health medication and to explore factors associated with initiation.

## METHODS

### Study design

The REporting of studies Conducted using Observational Routinely-collected health Data (RECORD) statement has been used for reporting this retrospective cohort study [17].

### Setting and data source

Data were collected as part of a larger study from 44 general practices (GP) in the Republic of Ireland using Socrates patient management software (www.socrates.ie) between 2011 and 2017 [18]. Data included demographic, clinical, prescribing and hospitalisation records of patients who were ≥65 years at the date of data extraction (2017). Ethical approval was obtained from the Irish College of General Practitioners.

### Participants

Patients were selected for inclusion in analysis if they had any records of prescriptions, hospitalisations or GP consultations over the study period (2011-2017) and were not prescribed any bone-health medication within 12 months of their first record. Patients were excluded if they had less than 12-months’ data.

Prescriptions of bone-health medication were defined as bisphosphonates, denosumab, raloxifene, parathyroid hormone, strontium ranelate or calcitonin and identified from either GP prescription records or hospitalisation discharge summaries. WHO Anatomical Therapeutic Chemical (ATC) classification codes (www.whocc.no/atc_ddd_index) were used to identify GP prescriptions (Appendix Table 1). Medications in discharge summaries were identified with free-text search terms of trade and generic names based on the Irish Health Products Regulatory Authority (www.hpra.ie) database (Appendix Table 1).

**Table 1.**
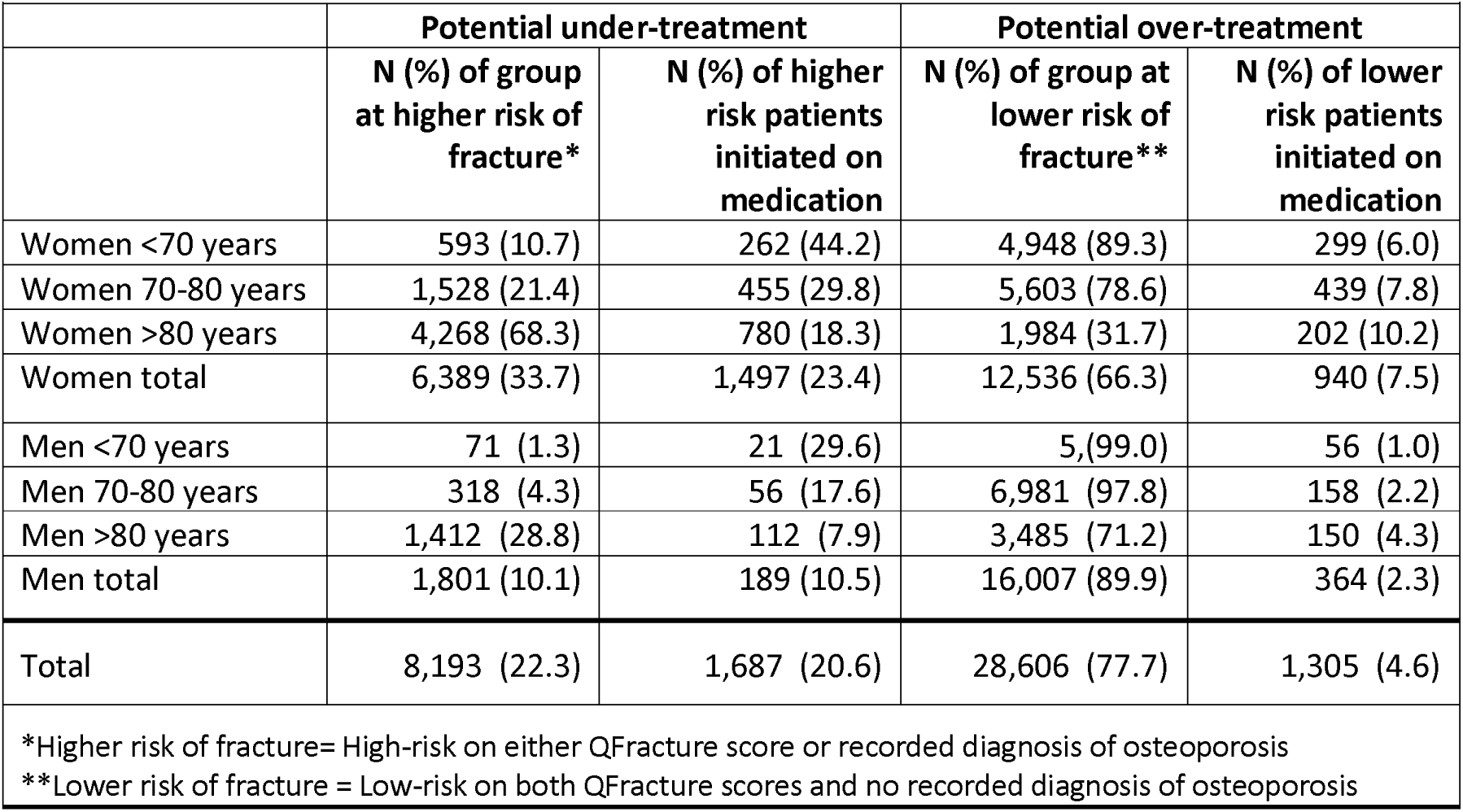
Patients by fracture-risk age, sex and initiated group.

### Outcome: Bone-health medication initiation

Bone-health medication initiation was defined as receiving a bone-health prescription, where no such prescription was recorded in the preceding 12 months. The initiation date was defined as the “index time-point”. Patients with no bone-health medication initiation were defined as a comparator group, and a random “index time-point” assigned per patient after the initial 12-month period.

### Exposures

Exposures were defined in the pre index time-point period and included age, sex, osteoporosis diagnosis, a calcium/vitamin D prescription, health cover type and fracture risk factors. Health cover type was grouped into three categories based on whether patients pay at point-of-care: General Medical Services scheme (GMS, covering GP care, hospitalisations and medications), Doctor Visit Card (DVC, covering GP care only), and Private.

The presence of 17 fracture risk factors (conditions and prescriptions) based on the QFracture tool [16] was identified for each patient prior to the index time-point. The QFracture tool was chosen as it is particularly suited to use in primary care databases in the UK and Ireland, has shown good accuracy for predicting fracture-risk, and the 2012 score is available for open source use [4,19,20]. The presence of each condition was defined from consultations, GP prescriptions and hospitalisation records using previously validated ICD-10-AM, ICPC-2, ATC codes and free-text phrases (Appendix Table 2). Prescriptions were defined from ATC codes in GP prescriptions and free-text phrases from hospitalisation records (Appendix Table 2). Excessive alcohol use was defined dichotomously based diagnostic indicators related to alcohol dependency.

Five-year QFracture hip fracture risk and all-fracture risk were calculated based on open source formulae (www.qfracture.org). As data was not available for ethnicity, smoking status, body mass index (BMI), residence, or family history of osteoporosis, these variables were set to the default value in the calculation (community-dwelling, white, non-smoker, no osteoporosis family history and BMI missing) [16]. Patients were categorised as “high-risk” of fracture (top 10^th^ percentile) based on QFracture cut-off values derived by Dagan et al (4.0% for hip fracture and 6.7% for all fractures) [20].

### Co-variates

Co-variates in the analysis included total pre index time-point observation in days, number of GP consultations and number of hospitalisations. The number of unique prescribed medications was calculated per person for 12-months prior to the index time-point.

### Statistical analysis

The number and proportion of patients with either an osteoporosis diagnosis or defined as high-risk on QFracture scores were presented by age, sex and bone-health initiation status to describe potential under and over treatment. Demographic and clinical variables were described for initiation and non-initiation groups and compared using t-tests, chi^2^ and Wilcoxon rank-sum tests as appropriate.

Generalised linear model (GLM) regression using Poisson distribution was conducted to assess whether QFracture high 5-year risk of all fractures and hip fractures were associated with initiation of bone-health medication univariablely and adjusting for osteoporosis diagnosis, health cover type and co-variates. A pre-planned subgroup analysis was conducted to assess this relationship in males and females separately and a sensitivity analysis was conducted using continuous QFracture score. GLM regression was conducted to assess whether each fracture risk factor was associated with initiation of bone-health medication, adjusting for osteoporosis diagnosis, age, sex, health cover type and co-variates.

Relative risks were calculated and 95% confidence intervals (CIs) were adjusted for clustering of patients within GP practices. Stata 16 (StataCorp. 2019) was used for analyses and statistical significance was assumed at p<0.05.

## RESULTS

### Participants

Among 36,799 patients (51.4% female) naïve for bone-health medication included in the analysis (Figure 1), mean age at the index time-point was 75.4 (SD=8.4) years. Overall, 2,992 patients were initiated on bone-health medication during the study. In 62% this involved oral bisphosphonates, 33% initiated denosumab, and 3% initiated strontium. The remaining 2% initiated parathyroid hormone, raloxifene, bisphosphonate infusion or calcitonin. Most new-initiations (89%) were detected from GP prescriptions while 11% were based on hospital discharge summaries. Denosumab accounted for 20% of hospital initiations and 35% of GP initiations.

**Figure 1.**
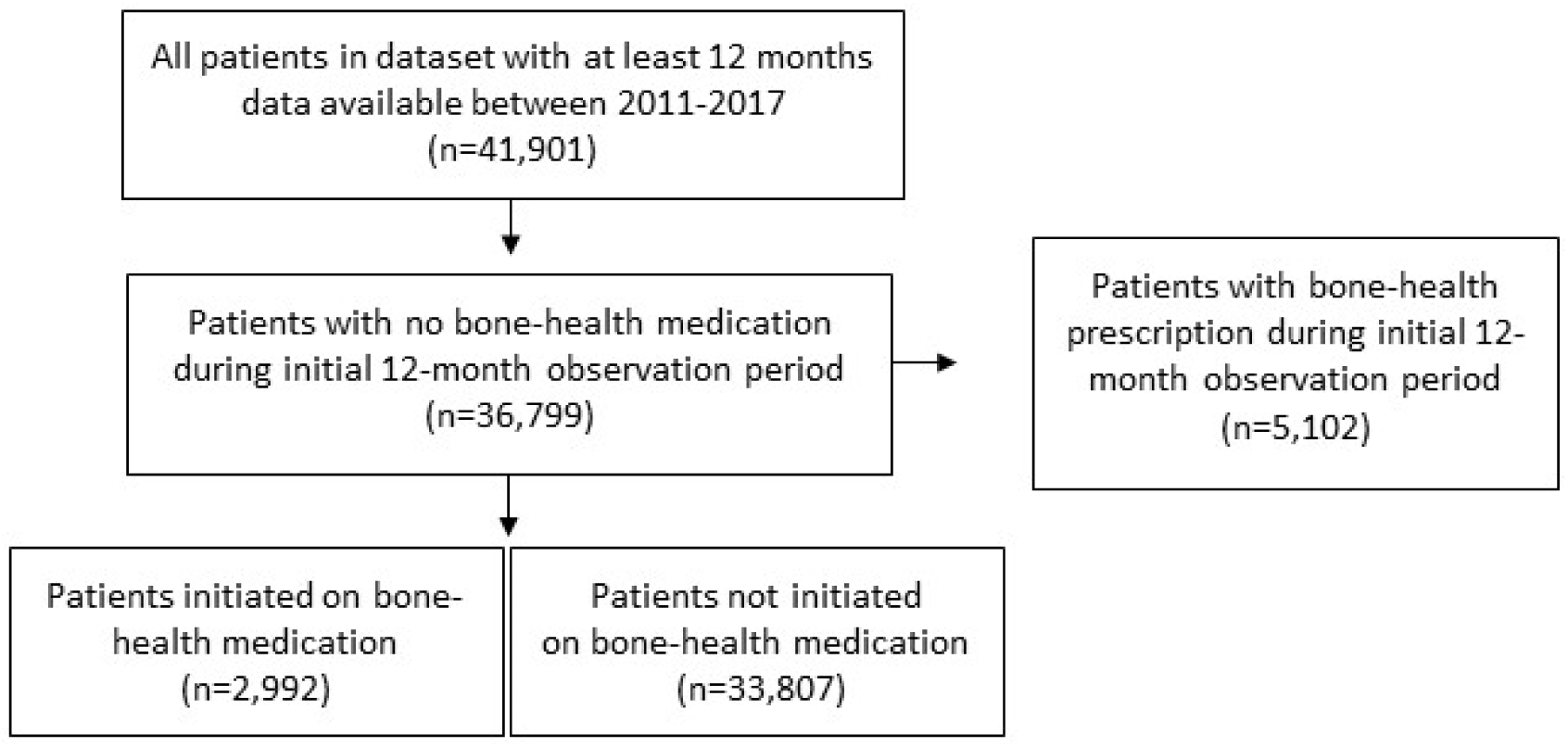
Flow-diagram of selected participants.

Almost 6% of patients (n=2,053) had an osteoporosis diagnosis but less than half of them (n=1,004; 49%) were initiated on bone-health medication. Furthermore, 22% of all patients (n=8,193) were found to either have osteoporosis or be high fracture-risk based on QFracture. Only 21% of them (n=1,687) were initiated on medication. Of 28,606 patients defined as lower fracture-risk with no osteoporosis diagnosis recorded, 5% (n=1,305) were initiated on medication. Table 1 shows this breakdown by sex and age-group.

### Factors associated with initiation of bone-health medication

Those initiated on bone-health medication were more likely to be female, have an osteoporosis diagnosis or GMS/DVC scheme coverage, and were older by 2.2 years on average (Table 2). Over 80% of those initiated had a pre-initiation calcium/vitamin D prescription in comparison to 18% of those not initiated.

**Table 2.** Demographics and healthcare factors by bone-health medication initiation status.

|  |  |  |  |
| --- | --- | --- | --- |
|  | Initiated (n=2,992) | Not Initiated (n=33,807) | p-value |
| Age at index time-point (mean (SD)) | 77.4 (8.2) | 75.2 (8.5) | <0.01 |
| Female sex (%) | 2437 (81.5) | 16488 (48.8) | <0.01 |
| Osteoporosis diagnosis (n (%)) | 1,004 (33.6) | 1,049 (3.1) | <0.01 |
| On Calcium/ Vitamin D (n (%)) | 2453 (82.0) | 5977 (17.7) | <0.01 |
| Health Cover (n (%)): |  |  | <0.01 |
| Private or Other | 537 (18.0%) | 10,189 (30.2%) |  |
| General Medical Services card | 2,138 (71.5%) | 20,554 (60.8%) |  |
| Doctor Visit Card | 316 (10.6%) | 3,036 (9%) |  |
| Observation time in days (mean (SD)) | 817 (587) | 706 (597) | <0.01 |
| Number of hospitalisations (median (IQR)) | 0.9 (2.4) | 0.5 (1.7) | <0.01 |
| Number of consultations (median (IQR)) | 33 (18-59) | 22 (10-41) | <0.01 |
| Number medications (median (IQR)) | 9 (5-14) | 7 (3-11) | <0.01 |

Independently of osteoporosis, health cover and co-variates, QFracture high 5-year risk for hip fractures (IRR=1.33, 95% CI=1.17-1.50, p<0.01) and all fractures (IRR=1.30, 95% CI=1.17-1.44, p<0.01) were associated with higher rates of medication initiation (Table 3). This relationship remained statistically significant in the subgroup analysis for males and hip fracture. Results were similar in analysis of continuous QFracture scores and initiation (Appendix Table 3), however all-fracture risk was also associated with initiation in males.

**Table 3.** Association between fracture-risk and bone-health medication initiation in all patients and subgroups of females and males.

|  |  |  |  |  |  |  |  |  |
| --- | --- | --- | --- | --- | --- | --- | --- | --- |
| QFracture 5-year risk | Initiated (n=2,992) | Not Initiated (n=33,807) | Univariable |  |  | Multivariable |  |  |
| All fragility fractures |  |  | IRR | 95% CI* | p-value | IRR** | 95% CI* | p-value |
| High | 864 (28.9%) | 5,186 (15.4%) | 2.06 | (1.85 to 2.30) | <0.01 | 1.33 | (1.17 to 1.50) | <0.01 |
| Low | 2126 (71.1%) | 28,557 (84.6%) |  |  |  |  |  |  |
| **Hip fractures** |  |  |  |  |  |  |  |  |
| High | 808 (27.0%) | 5,064 (15.0%) | 1.95 | (1.74 to 2.17) | <0.01 | 1.30 | 1.17 to 1.44 | <0.01 |
| Low | 2,182 (73.0%) | 28,679 (85.0%) |  |  |  |  |  |  |
| **Sub-group analysis by sex:** |  |  |  |  |  |  |  |  |
| QFracture 5-year risk in Females | Initiated females (n=2,437) | Not Initiated females (n=16,488) | Univariable |  |  | Multivariable |  |  |
| All fragility fractures |  |  | IRR | 95% CI* | p-value | IRR** | 95% CI* | p-value |
| High | 787 (32.3%) | 4,030 (24.4%) | 1.40 | (1.24 to 1.58) | <0.01 | 1.08 | (0.96 to 1.21) | 0.19 |
| Low | 1,650 (67.7%) | 12,458 (75.6%) |  |  |  |  |  |  |
| **Hip fractures** |  |  |  |  |  |  |  |  |
| High | 711 (29.2%) | 3,598 (21.8%) | 1.40 | (1.24 to 1.57) | <0.01 | 1.10 | (0.99 to 1.22) | 0.06 |
| Low | 1,726 (70.8%) | 12,890 (78.2%) |  |  |  |  |  |  |
| QFracture 5-year risk in Males | Initiated males (n=553) | Not Initiated males (n=17,255) |  |  |  |  |  |  |
| All fragility fractures |  |  |  |  |  |  |  |  |
| High | 77 (13.9%) | 1,156 (6.7%) | 2.17 | (1.74 to 2.71) | <0.01 | 1.19 | 0.97 to 1.46 | 0.10 |
| Low | 476 (86.1%) | 16,099 (93.3%) |  |  |  |  |  |  |
| **Hip fractures** |  |  |  |  |  |  |  |  |
| High | 97 (17.5%) | 1,466 (8.5%) | 2.21 | (1.81 to 2.70) | <0.01 | 1.26 | 1.07 to 1.50 | <0.01 |
| Low | 456 (82.5%) | 15,789 (91.5%) |  |  |  |  |  |  |
| *95% CIs adjusted by General Practice clusters<br>**IRR adjusted for osteoporosis diagnosis, health cover, observation time, number of hospitalisations (in 4 categories), number of consultations (in quartiles), number of medications in prior 12 months (in quartiles). |  |  |  |  |  |  |  |  |

Independently of age, sex, osteoporosis, health cover and co-variates, fracture history (IRR=1.8, 95% CI=1.6-2.0), Rheumatoid Arthritis/SLE diagnoses (IRR=1.8, 95% CI=1.5-2.2), and corticosteroid use (IRR=1.6, 95% CI=1.5-1.8) were associated with medication initiation (Table 4). Diabetes (IRR=0.7, 95% CI=0.6-0.7), chronic liver disease (IRR=0.7, 95% CI=0.5-0.8), chronic kidney disease (IRR=0.8, 95% CI=0.7-1.0), cardiovascular disease (IRR=0.9, (95% CI=0.8-0.9) and oestrogen-only hormone replacement therapy use (IRR=0.7, 95% CI=0.6-0.9) had an inverse association with medication initiation.

**Table 4.** Association between fracture risk factors and bone-health medication initiation.

| Conditions/ prescriptions | Initiated<br>(n=2,992) | Not Initiated<br>(n=33,807) | IRR* | 95% CI** | p-value |
| --- | --- | --- | --- | --- | --- |
|  | (n (% of initiation group)) |  |  |  |  |
| Asthma or COPD | 954 (31.9) | 8,276 (24.5) | 1.1 | (1.0 to 1.1) | 0.16 |
| On antidepressants | 755 (25.2) | 6,145 (18.2) | 1.0 | (0.9 to 1.1) | 0.91 |
| On corticosteroids | 582 (19.5) | 2,772 (8.2) | 1.6 | (1.5 to 1.8) | <0.01 |
| Cardiovascular disease | 527 (17.6) | 5,817 (17.2) | 0.9 | (0.8 to 0.9) | <0.01 |
| Cancer | 462 (15.4) | 4,069 (12.0) | 1.0 | (0.9 to 1.1) | 0.84 |
| Fragility fracture | 320 (10.7) | 814 (2.4) | 1.8 | (1.6 to 2.0) | <0.01 |
| Diabetes mellitus | 274 (9.2) | 4,744 (14.0) | 0.7 | (0.6 to 0.7) | <0.01 |
| Dementia | 205 (6.9) | 1,930 (5.7) | 1.0 | (0.9 to 1.1) | 0.68 |
| Epilepsy | 158 (5.3) | 1,286 (3.8) | 1.1 | (0.9 to 1.3) | 0.41 |
| Chronic kidney disease | 148 (5.0) | 1,314 (3.9) | 0.8 | (0.7 to 1.0) | 0.04 |
| RA or SLE | 140 (4.7) | 447 (1.3) | 1.8 | (1.5 to 2.2) | <0.01 |
| History of falls | 62 (2.1) | 360 (1.1) | 1.1 | (0.9 to 1.4) | 0.46 |
| Parkinson's disease | 110 (3.7) | 974 (2.9) | 1.1 | (1.0 to 1.4) | 0.14 |
| Endocrine disorders | 96 (3.2) | 537 (1.6) | 1.1 | (0.9 to 1.2) | 0.50 |
| Diseases of malabsorption | 40 (1.3) | 263 (0.8) | 1.1 | (0.8 to 1.4) | 0.50 |
| Oestrogen only HRT | 45 (1.5) | 402 (1.2) | 0.7 | (0.6 to 0.9) | <0.01 |
| Chronic liver disease | 34 (1.1) | 423 (1.3) | 0.7 | (0.5 to 0.8) | <0.01 |
| Alcohol dependency | 38 (1.3) | 473 (1.4) | 1.0 | (0.9 to 1.2) | 0.52 |

COPD=Chronic Obstructive Pulmonary Disease; RA=Rheumatoid Arthritis; SLE=Systemic Lupus Erythematosus; HRT=Hormone Replacement Therapy
\*IRR adjusted for osteoporosis diagnosis, sex, age, health cover type, pre-initiation observation time, number of hospitalisations (4 categories) and number of consultations (quartiles), number of medications in prior 12 months (quartiles).
\*\*95% CIs adjusted by General Practice clusters

## DISCUSSION

This study of bone-health naïve older adults attending primary care practices in Ireland found that only 23% of women and 11% of men at high-risk of fragility fracture were initiated on bone-health medication. In comparison, 5% of those not defined as high fracture-risk were initiated on medication.

Patients who were newly initiated on bone-health medication were older, more likely to be female and to have state-funded health cover. While QFracture scores were associated with higher rates of medication initiation, this relationship was not observed in the female subgroup, potentially suggesting that female sex is the main factor considered by clinicians.

Independently of age, sex and osteoporosis diagnosis, specific fracture risk factors associated with medication initiation included fracture history, corticosteroid use, and rheumatoid arthritis/SLE diagnosis. In contrast, patients with diabetes, chronic liver or kidney disease, or cardiovascular disease were less likely to be initiated.

The findings of this study are broadly in line with previous research in Ireland and internationally suggesting that fracture risk is under-identified and under-treated in older adults in primary care [9,12,13]. Potential under-treatment was particularly notable in men >80 years old with only 8% of those at higher fracture-risk being initiated on medication. Irish GPs have previously reported having good awareness of female osteoporosis but the condition in men is less recognised [21]. While men have lower hip fracture incidence, those who do fracture are younger with more co-morbidities and experience worse outcomes including higher mortality [22]. Over-looking the assessment of fracture risk and primary and secondary fracture prevention in this vulnerable population could therefore have significant clinical consequences.

Factors identified in this study as most strongly associated with medication initiation are those most frequently described in guidelines, namely female sex, osteoporosis, fragility fracture history and corticosteroid use [4-7,10]. These factors have also been reported by Irish GPs as influencing their decision to consider an osteoporosis diagnosis [21]. It is not known to what extent Irish GPs take other clinical factors into account or use prediction tools including QFracture and FRAX, and warrants further research.

Lower levels of treatment in patients with specific medical conditions, including diabetes, kidney, liver and cardiovascular disease, may reflect previous findings that GP concerns about medication side effects and burden can influence decision-making [9,13,14]. The benefit of taking an additional medicine is likely to reduce in the presence of existing polypharmacy [23].This is important, as older adults with the highest fracture-related mortality rates are more likely to have multi-morbidity. This may explain to some degree potential under-treatment observed in the oldest age group.

There may be understandable clinical dilemmas for GPs deciding whether to prioritise pharmacological fracture-prevention in the presence of particular medical conditions. Common adverse effects that should influence decision-making include upper gastrointestinal events for bisphosphonates [7] and infections for denosumab [24]. While oesophageal symptoms are common in liver disease and stroke, these conditions also contribute significantly to osteoporosis and GPs could consider alternative medicine formulations in these circumstances [4,16,25]. Severe kidney impairment may complicate osteoporosis treatment due to calcium homeostasis disturbances and reduced drug excretion, and specialist referrals may be indicated for patients with high fracture risk [4,26]. Bisphosphonates or denosumab have not been found to increase the risk of cardiovascular events so the reasoning behind a lower prescription in patients with these conditions as found in this study is not clear [7,27,28]. Small observational studies have linked diabetes to medication-related jaw osteonecrosis, however this has been refuted in higher quality research [29]. Osteoporosis guidelines should provide clear advice regarding risk-benefit decisions in the presence of medical conditions, providing information on testing required and in which cases referral to specialists are indicated [4]. GPs may require further support and education in this area.

We were unable to consider lifestyle measures in osteoporosis management including exercise, smoking cessation, alcohol reduction and falls-prevention [4]. We did however find that over two-thirds of patients prescribed calcium and/or vitamin D supplements were not initiated on bone-health medication. While adequate dietary intake of these substances is important for maintaining BMD in older adults and as an adjunct to pharmacotherapy, GPs should be aware that supplementation without bone-health medication prescription has not been shown to reduce fracture incidence in those at high-risk [4].

### Strengths and limitations

This study includes a large cohort of older adults treated in primary care. We ascertained prescribing and fracture-risk factors from multiple sources (GP prescriptions, consultations and hospitalisation discharge summaries). Using data available, we were unable to include information on residence, osteoporosis family history, BMI, and smoking status in calculating QFracture scores. In addition, as falls were identified based only on hospitalisation records, they are likely under-coded resulting in a 2% prevalence in comparison to the 20% that would be expected [12]. These factors likely led to an underestimation of fracture-risk, suggesting under-treatment may actually be higher. To overcome this bias when assessing the relationship between QFracture score and medication initiation, a sensitivity analysis of the continuous score was conducted, with consistent results.

Using GP prescribing records, we could not determine if patients received prescriptions from consultant geriatricians or fracture liaison services, however in the Irish setting patients would likely return to their GP for repeat prescriptions. We were also unable to fully assess reasons for non-initiation of medication that may have been clinically appropriate and to what extent it reflects patient preferences.

### Clinical implications

Interventions to improve appropriate initiation of bone-health medication in patients with high fracture-risk require investigation in the Irish primary care setting. International studies have shown benefits for complex interventions that include patient and practitioner education with feedback about test results/medication and encouragement for patients to follow-up with their practitioners [30]. This study’s findings suggest that GPs may need particular support in making risk/benefit decisions about prioritising pharmacological treatment for fracture prevention, especially in adults >80 years old with medical co-morbidities.

## CONCLUSION

While there is a higher rate of bone-health medication initiation among older patients in primary care in Ireland who are at higher fracture-risk, there remains much potential under-treatment. This is particularly true among oldest patients and those with chronic medical co-morbidities. Future research should assess reasons for non-prescription of bone-health medication in Ireland, including patient and GP concerns, and interventions to support appropriate prescribing in this setting.

## Data Availability

No provision for data sharing was included in initial ethical approval.

## Notes

### Competing Interest Statement

The authors have declared no competing interest.

### Funding Statement

Support was received from the Health Research Board (HRB) in Ireland through grant no. HRC/2014/1 (TF) and through the RCSI Research Summer School 2019 (MEW and MN).

### Author Declarations

Ethical approval was obtained from the Irish College of General Practitioners.

