## Appendix for "Factors associated with initiation of bone-health medication among older adults in primary care in Ireland"

| Appendix Table 1. Bone-health medication definitions | | |
|---|---|---|
| **Medication group** | **ATC codes** | **Free-text search terms** |
| Bisphosphonates (Oral) | M05BA01 M05BA02 M05BA03 M05BA04 M05BA05 M05BA06 M05BA07 M05BB | aclasta actonel alendromax alendronic aredia binosto bonapenya bonasol bondenza bondronat bonefos bonefurbit bonviva clasteon destara didronel pamidronate fosalen fosamax fostepor fostolin iasibon ibandronic ibandronate kefort loron optinate osbonelle osteomel ridate risedronate riseseus risonate risontel romax teboneva tevanate zerlinda adrovance alendronate didronel fosavance optinate ridate vantavo etidronic etidronate clodronic clodronate pamidronic tiludronic tiludronate alendronaate aledronic alenronic risedronic |
| Bisphosphonates (Infusion) | M05BA08 | zoledronic zometa zoledronate |
| Denosumab | M05BX04 | denosumab prolia xgeva |
| Raloxifene | G03XC01 | raloxiep raloxifene |
| Parathyroid hormone | H05AA | parathyroid forsteo movymia natpar preotact terrosa tetridar teriparatide |
| Strontium | M05BX03 M05BX53 | osseor protelos strontium |
| Calcitonin | H05BA | calsynar forcaltonin miacalcic miakaril ostulex calcitonin elcatonin |

| **Appendix Table 2. Fracture risk factor and other exposure condition and prescription definitions*** | | | | |
|---|---|---|---|---|
| **Condition** | **ICD-10** | **ICPC-2** | **ATC** | **Free-text search terms** |
| Osteoporosis** | M80- M82 | L95 | | osteop paget |
| Asthma or COPD | J40-J46 J68.4 | R95 R96 | R03AC R03AH R03AK R03AL R03B R03C R03DA R03DB R03DC01 R03DC02 R03DC03 R03DX | asthma copd "chronic obstructive pulmonary disease" "emphysema" "chronic bronchitis" |
| Cardiovascular Disease (including heart attack, angina, stroke and transient ischaemic attack) | I20 I21 I23 I24.0 I24.8 I24.9 I25.0 I25.1 I25.8 I25.9 I70 G45 G46 H34.0 I60 I61 I62 I63 I64 I65 I66 I67 I68 I69 H34.1 | K74 K75 K76 K89 K90 K91 | C01DA02 C01DA04 C01DA05 C01DA07 C01DA08 C01DA09 C01DA13 C01DA14 C01DX16 C08EX02 | stemi "myocardial infarct" angina "coronary disease" " ihd " " cad " "coronary artery disease" "cerebral infarct" cva stroke "ischaemic attack" "ischemic attack" cerebrovascular " tia " |
| Cancer | C00-C97, D00-D09 | A79 B72 D74 D75 D76 D77 F74 H75 K72 L71 N74 R84 R85 S77 T71 U75 U76 U77 X75 X76 X77 Y77 Y78 | L01A L01B L01C L01D L01XA L01XB L01XC L01XD L01XE L01XX01 L01XX03 - L01XX41 | cancer carcinoma neoplasm tumor tumour malignan metastatic "breast ca" "lung ca" "prostate ca" |
| Fragility Fracture | M48.4 M48.5 S22.0 S22.1 S32.0 S42 S52 S72 | L72 L75 L76 | | colles "wrist fracture" "distal radius fracture" "compression fracture" "fatigue fracture""neck of femur" "hip #" "nof" "osteoporosis with pathological fracture" "fracture of lower end of radius" "fracture of humerus" "vertebral collapse" "hip fracture" "femoral fracture" |

| | | | | |
|---|---|---|---|---|
| Diabetes Mellitus | E10 E11 E13 E14 | T89 T90 | A10A A10BA01-A10BA03 A10BB01- A10BB12 A10BB31 A10BC01 A10BD01 - A10BD25 A10BF01- A10BF03 A10BG01- A10BG03 A10BH01- A10BH07 A10BH51 A10BH52 A10BJ01 - A10BJ06 A10BK02 A10BK04 - A10BK06 A10BX01-A10BX08 A10BX10 A10BX11 A10BX13 A10BX14 A10BK01 A10BK03 | diabetes diabetic " dm" |
| Type 2 | E11 | T90 | A10BA01-A10BA03 A10BB01- A10BB12 A10BB31 A10BC01 A10BD01 - A10BD25 A10BF01- A10BF03 A10BG01- A10BG03 A10BH01- A10BH07 A10BH51 A10BH52 A10BJ01 - A10BJ06 A10BK02 A10BK04 -A10BK06 A10BX01-A10BX08 A10BX10 A10BX11 A10BX13 A10BX14 A10BK01 A10BK03 | t2dm dmII "type 2 dm" |
| Type 1 | E10 | T89 | A10A | t1dm "type 1 dm" |
| Dementia | F00 F01 F02 F03 F05.1 G30 G31.1 | P70 | N06DA02 N06DA03 N06DA04 N06DX01 | dementia alzheimer |
| Epilepsy | G40 G41 | N88 | N03AB N03AC N03AD N03AD N03AE N03AF N03AG N03AX03 N03AX07 N03AX09 N03AX10 N03AX11 N03AX13 N03AX14 N03AX15 N03AX17 N03AX18 N03AX19 N03AX21 N03AX22 N03AX23 N03AX24 N03AX30 | epilepsy epilepticus seizure convulsion |
| Chronic Kidney Disease (stage 4 or 5) | N00 - N23 N25 I12.0 I13.1 Z49.0 Z49.1 Z49.2 Z94.0 Z99.2 | U88 U95 U99 | A11CC01 A11CC02 A11CC03 A11CC04 B03XA01 B03XA02 B03XA03 V03AE02 V03AE03 V03AE | pyelonephritis "renal failure" "kidney failure" "kidney disease" " ckd " eskd "urinary calculus" "renal disease" "renal failure" "kidney transplant" |

| | | | | |
|---|---|---|---|---|
| Rheumatoid arthritis/ Systemic Lupus Erythema | M05 M06 M31.5 M32 M33 M34 M35.1 M35.3 M36 | L88 | | rheumatoid polymyalgia " ra " " sle " lupus |
| History of falls | W0 W1 R29.81 R29.6 R55 | | | fall fell vasovag " syncope" |
| Parkinson’s Disease | G20-G22 | N87 | N04A N04BA N04BB N04BC N04BD N04BX01 N04BX02 | parkinson |
| Endocrine Disorders | E05 E21 E24 | T81 T85 | H03BA02 H03BB01 | hyperthyroid thyrotox hyperparathy cushing |
| Diseases of Malabsorption (Including Chron’s disease and ulcerative colitis) | K50 K51 K90 | D94 | | chrons chron's ibd "ulcerative colitis" "inflammatory bowel" celiac coeliac steatorrhea "blind loop" |
| Chronic Liver Disease | I85.0 I85.9 I98.2 I98.3 K65.0 K65.8 K65.9 K67.0 K67.1-K67.3 K67.8 K70.3 K74.3-K74.6 K76.7 K93.0 R18 | D72 D97 | | cirrhosis "liver disease" |
| Alcohol dependency | E52 F10 G62.1 I42.6 K29.2 K70.0 K70.3 K70.9 T51 Z50.2 Z71.4 Z72.1 | P16 P15 | N07BB | alcoholic "alcohol related" "alcohol-related" |

| Appendix Table 2 continued. Fracture risk factor and other exposure condition and prescription definitions* | | |
|---|---|---|
| **Prescriptions** | **ATC** | **Free-text search terms** |
| Prescription of calcium/ vitamin D** | A12A, A11CC, A11CB | actonel altavita "at 10" bellcalcid bocatriol cacit cadelius calcichew calciforte calcijex calciup cal-d-vita calfovit caltrate calvidin crampex desunin everose fultium-d3 "halibut liver oil" ideos kalcipos-d "one-alpha" osteocur osteofos ostram osvaren rocaltrol sandocal sapvit-d3 silkis teboneva thorens calcium ergocalciferol dihydrotachysterol alfacalcidol calcitriol colecalciferol calcifediol "vitamin a and d" "vitamin d" cholecalciferol calciche calciferol cholecalciferol calcihew calcidol alfacalcidiol calcitrol cholecalcierol |
| Corticosteroids (>2 scripts) | H02AB H02BX M01BA | adcortyl alkindi betnelan betnesol calcort decadron deltacortril depo-medrone dexliq dexsol dilacort hydrocortone lodotra medrone neofordex oradexon pevanti plenadren prednesol solu-cortef solu-medrone betamethasone cloprednol cortisone cortivazol deflazacort dexamethasone fluocortolone hydrocortisone meprednisone methylprednisolone paramethasone prednisolone prednisone prednylidene rimexolone triamcinolone prenisolone prednisalone prednislone prednisolon |
| Antidepressants (>2 scripts) | N06A | agomelatine alaproclate amineptine amiti amitr amotrip amoxapine amylt amyt anafranil ariclaim asendis astillin bellcital bellsert bellzac bexmirt bexzis bezxis bifemelane biozac brintellix bupropion butriptyline camcolit ciprager cipralam cipraml ciprapine cipreger ciprimil citala citalipram citalo citalpra citola citrol citropram clomipramin concordin cymbalta cytalopram depreger Desipramine desvenlafaxine dibenzepin dilox dimetacrine doluxetin doselupin dosulepin dothep doxepin doxipin dulox duloxetine dutonin ecitalopram edronax efaxil efex effexor escial esciprex escital escitalopram escitalpro escitomar escitotab escivriens esitalopram etalopro etoperidone faverin faxin floxetin fluox fluzac gama gepirone geramil gerozac gerzac imipramine iprindole iproclozide iproniazide ireven isocarboxazid lentizol lexapro lexepro lexpro lithium lofepramine lofipramine lofpramine loxentia lusert lustr lustral luvoxamine mainserin majoven manerix maprotiline medifoxamine melitracen meloxat mianserin miansiren milnacipran minaprine mirap mirat mirt mirtaz mirtazapine mirtazipine mirzaten moclo moclobemide moclobemide moclobimide molco molipaxin molopaxin molypaxin nefazodone nialamide nomifensine nortr nortriptyline norzac olena opipramol oxaflozane oxitriptan pantox parnate parocetan paroser parox paroxetin paroxetine paroxitine paxt phenelzine phenelzine pivagabine priadel proth prothiaden protriptyline prozac prozamel prozatan prozit prozmel pyrox quinupramine reboxetine rethera scippa seretral serimel serlan serox seroxat serta sertarlin sertr sertraline sertraniche setarlin setralin setraline setrol sinequan surmontil symbal tardcaps tazamel thymanax tianeptine tofranil toloxatone tolvon tonpular tranylcypromine trazodone trazondone trazone trimipramine triptizol trymip tryptizol tryptophan valadoxan valdex valdoxan vedixal velexor venel venex venfax venifax venlablue venlafaxine venlafex venlalic venlift venlofex vilazodone viloxazine vortioxetine xepin xeristar zimeldine zismirt zispin zistap zyban |
| Oestrogen only HRT (>2 scripts) | G03CA | aerodiol alora binovum blissel cathate climara climaval dermestril divigel elleste-solo epiestrol estradot estramon estreva estrofem evorel fematab fematrix imvaggis lenzetto menorest oesclim oestrogel ortho-gynest ovestin ovysmen premarin sayana vagifem chlorotrianisene estradiol estriol estrogen estrone ethinylestradiol promestriene |

*The codes presented were derived based on previous international literature [1-8] and adapted for the Irish clinical setting. They were validated in the full dataset by comparing the prevalence of co-morbidities found in the dataset to other research among older general practice populations internationally and the QFracture derivation and validation studies [1, 9-16]
** Not included in QFracture Score

| Appendix Table 3. Sensitivity analysis - Association between linear Qfracture risk score and bone-health medication initiation in all patients and subgroups of females and males | | | | | | | | |
|---|---|---|---|---|---|---|---|---|
| **QFracture 5-year risk** | **Initiated (n=2,992)** | **Not Initiated (n=33,807)** | **Univariable** | | | **Multivariable** | | |
| | | | **IRR** | **95% CI*** | **p-value** | **IRR**** | **95% CI*** | **p-value** |
| **All fragility fracture score (mean (SD))** | 5.8 (4.8) | 3.9 (4.3) | 1.05 | (1.05 to 1.06) | <0.01 | 1.03 | (1.02 to 1.04) | <0.01 |
| **Hip fracture score (mean (SD))** | 3.6 (4.9) | 2.3 (4.0) | 1.04 | (1.03 to 1.04) | <0.01 | 1.02 | (1.01 to 1.02) | <0.01 |
| **Sub-group analysis in by sex:** | | | | | | | | |
| **QFracture 5-year risk in females** | **Initiated females (n=2,437)** | **Not Initiated females (n=16,488)** | **Univariable** | | | **Multivariable** | | |
| | | | **IRR** | **95% CI*** | **p-value** | **IRR**** | **95% CI*** | **p-value** |
| **All fragility fracture score (mean (SD))** | 6.2 (4.8) | 5.4 (4.4) | 1.03 | (1.02 to 1.04) | <0.01 | 1.01 | (1.00 to 1.02) | 0.22 |
| **Hip fracture score (mean (SD))** | 3.7 (5.0) | 3.0 (4.4) | 1.02 | (1.01 to 1.03) | <0.01 | 1.01 | (1.00 to 1.02) | 0.07 |
| **QFracture 5-year risk in males** | **Initiated males (n=553)** | **Not Initiated males (n=17,255)** | | | | | | |
| **All fragility fracture score (mean (SD))** | 4.1 (4.7) | 2.4 (3.6) | 1.05 | (1.04 to 1.06) | <0.01 | 1.02 | (1.01 to 1.03) | <0.01 |
| **Hip fracture score (mean (SD))** | 2.9 (4.6) | 1.6 (3.4) | 1.04 | (1.03 to 1.05) | <0.01 | 1.02 | (1.01 to 1.03) | <0.01 |

* 95% Cis adjusted for General Practice clusters
*IRR adjusted for pre-initiation osteoporosis diagnosis, number of medications (in quartiles), health cover, pre-initiation observation time, number of hospitalisations (in 4 categories), number of consultations (in quartiles)
